# Modeling *Plasmodium falciparum* Diagnostic Test Sensitivity using Machine Learning with Histidine-Rich Protein 2 Variants

**DOI:** 10.1101/2020.05.27.20114785

**Authors:** Colby T. Ford, Gezahegn Alemayehu, Kayla Blackburn, Karen Lopez, Cheikh Cambel Dieng, Eugenia Lo, Lemu Golassa, Daniel Janies

## Abstract

Malaria, predominantly caused by *Plasmodium falciparum*, poses one of largest and most durable health threats in the world. Previously, simplistic regression-based models have been created to characterize malaria rapid diagnostic test performance, though these models often only include a couple genetic factors. Specifically, the Baker et al., 2005 model uses two types of particular repeats in histidine-rich protein 2 (PfHRP2) to describe a *P. falciparum* infection [1], though the efficacy of this model has waned over recent years due to genetic mutations in the parasite. In this work, we use a dataset of 102 *P. falciparum* PfHRP2 genetic sequences collected in Ethiopia and derived a larger set of motif repeat matches for use in generating a series of diagnostic machine learning models. Here we show that the usage of additional and different motif repeats proves effective in predicting infection. Furthermore, we use machine learning model explanability methods to highlight which of the repeat types are most important, thereby showcasing a novel methodology for identifying potential targets for future versions of rapid diagnostic tests.

## 1 Introduction

Malaria affects over 228 million people and resulted in 405,000 deaths in 2018 [2]. Genomics is beginning to bear fruit in abatement of malaria but presents analytical challenges due to the complexity of the disease and its components (human, *Plasmodium spp*., and vector mosquitoes).

In most developing countries, the detection of *Plasmodium falciparum* and diagnosis of malaria is often performed using simple rapid diagnostic tests (RDTs). Specifically, these tests are lateral flow immuno-chromatographic antigen detection tests that are similar in modality to common at-home pregnancy tests. These tests use dye-labeled antibodies to bind to a particular parasite antigen and display a line on a test strip if the antibodies bind to the antigen of interest [3]. If patients are properly diagnosed, *P. falciparum* infections may be treated using antimalarial drugs such as artemisinin or artemisinin combined therapies (ACTs). Unfortunately, the efficacy of RDTs and artemisinin treatment have diminished in some settings around world, specifically in locations where the deletion or mutation of the kelch domain–carrying protein *K13* gene are observed [4].

In 2005, Baker et al. published a simple linear regression-based model that purports to predict the detection sensitivity of RDTs using a small fraction of genetic sequence variants that code for histidine-rich protein 2 (PfHRP2) [1]. While with the data available at the time, the accuracy of the Baker model was high (87.5%), the explanation ability of the RDT sensitivity was low (R2 = 0.353). Enthusiasm for the Baker model has since diminished. In 2010, Baker et al. published a report in which they concluded that they can no longer correlate sequence variation and RDT failure with their model [5].

Nevertheless, there is no alternative to the Baker model and it is still in use. In this study, our hypothesis is that a model for understanding the relationship between RDT and sequence variation can be improved by using a larger set of genetic sequence variants. Our purpose is to use large datasets and machine learning methods to address the shortcomings in malaria diagnosis test sensitivity and to provide a novel approach to direct the development of future RDTs using PfHRP2. In this study, we analyze a collection of genetic data and metadata from 102 *P. falciparum* sequences collected from Ethiopia with the Baker model along with a sweep of other machine learning models that we generate.

Beyond simply training a better model using more sophisticated algorithms, our research focus is to allow for interpretable insights of the machine learning models to be derived from the “black box”. We have shown previous success in AI-driven explanations of gene expression underlying drug resistant strains of *Plasmodium falciparum* [6, 7]. We apply this model interpretability here to identify which types of histidine-rich repeats, present in PfHRP2, are most indicative of malaria test performance.

## 2 Materials and Methods

## 2.1 Data Collection

Blood samples and demographic data were collected from suspected malaria patients greater than five years of age in various health clinics during both the low and high transmission seasons in different regions of Assosa, Ethiopia. Specifically, this health facility-based cross-sectional study was conducted in febrile patients seeking malaria diagnosis at four selected health facilities: Assosa, Bambasi, Kurmuk and Sherkole from November to December 2018.

Microscopy and rapid diagnostic testing were performed within the health clinics, and drops of blood spotted on Whatman 3MM filter paper were kept in sealed pouches for later analyses. CareStart™ malaria combination RDTs (lot code 18H61 from Access Bio Ethiopia) were used to diagnose *P. falciparum* and to evaluate their performance against microscopy as a reference test.

The *P. falciparum* DNA concentration in dried blood spot samples was analyzed using real-time quantitative PCR (RT-PCR). The *P. falciparum* DNA was extracted using phosphate buffered saline, Saponin, and Chelex [8] and confirmed *P. falciparum* positive samples as those whose RT-PCR values were less than or equal to 37 [9]. The null hypothesis was that RDT testing and the detection of *P. falciparum* by RT-PCR will have a strong correlation (e.g., positive RDT samples will lead to positive RT-PCR and negative RDT samples will lead to negative RT-PCR). However, early findings have shown incongruence between the RDT results and RT-PCR [10].

Using the primers listed in Table 1, two amplicons were sequenced, including a 600 to 960-bp fragment for Pfhrp2 Exon 2 [1] and a 294 to 552-bp fragment for Pfhrp3 Exon 2 [5]. Each sample was sequenced once, in both forward and reverse directions to create a consensus sequence for each sample. Polymerase Chain Reaction (PCR) conditions for Pfhrp2 Exon 2 and Pfhrp3 Exon 2 are shown in Table 1. The DNA amplicon quality was observed by means of agarose gel electrophoresis and the bands visualized in a UV transilluminator. PCR products were cleaned with 10 units of Exonuclease I (Thermo Scientific) and 0.5 units of shrimp alkaline phosphatase (Affymetrix) at 37 °C for 1 h followed by a 15 min incubation at 65 °C to deactivate the enzymes. PCR products were sequenced with ABI BigDye Terminator v3.1 (Thermo Fisher Scientific) following the manufacturer’s protocol using the conditions of (1) 95 °C for 10 s, (2) 95 °C for 10 s, (3) 51 °C for 5 s, (4) 60 °C for 4 min, and (5) repeat steps 2-4 for 39 more cycles. The samples were cleaned using Sephadex G-50 (Sigma-Aldrich) medium in a filter plate and centrifuged in a vacufuge to decant.

**Table 1:**
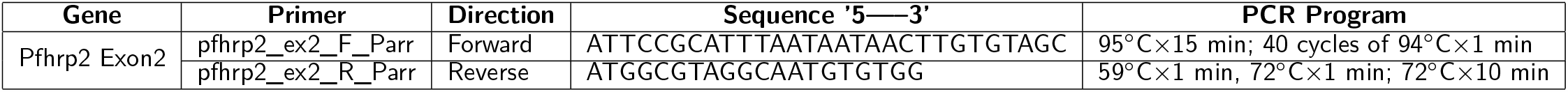
PCR Conditions and Primer Sequences from Parr et al., 2018 [11].

The samples were reconstituted with Hi-Di Formamide (Thermo Fisher Scientific) and the plates were placed on the ABI 3130 Sequencer. Sequence trace files from all samples and repeat samples were imported into CodonCode Aligner (CodonCode Corporation). The bases were called for each sample. The ends of the sequences were trimmed by the application when possible and manually when necessary. All sequences were examined and evaluated on both the forward and reverse strands, with manual base corrections and manual base calls occurring when necessary. This resulted in 102 usable sequences.

### 2.2 Data Preparation

All Pfhrp2 exon 2 nucleotide sequences were exported from CodonCode Aligner (CodonCode Corporation) and individually pasted into the ExPASy Translate tool (Swiss Institute of Bioinformatics Resource Portal). Both forward and reverse DNA strands were translated using the standard NCBI genetic code. The six reading frames of the amino acid sequence produced were examined. CodonCode’s default parameters were used for clipping the ends and a visual check was performed of each sequence to ensure base calls were correct, and trimmed further as needed.

For each nucleotide sequence, the amino acid sequence presenting the fewest number of stop codons was selected for further analysis. If two or more of the reading frames appeared to produce sequences with an equally minimal number of stop codons, the reading frame that produced a sequence exhibiting the previously recognized pattern in prior sequences was selected for further analysis. While most of the sequences had a clear, single best translation, 11 of the sequences required further editing. In these 11 sequences, the sequence portion before or after the stop codon which exhibited a pattern similar to prior sequences was used in analysis, while the portion of the sequence preceding or following the stop codon, which did not exhibit the recognized pattern, was discarded. Nucleotide sequence input into the ExPASy Translate Tool (Swiss Institute of Bioinformatics Resource Portal) was repeated and verified for accuracy of amino acid sequences. The verified sequences were compiled.

#### 2.2.1 Motif Search

A motif search was performed across 24 different types of histidine-based repeats. These repeat types, listed in Table 3, were originally defined by Baker et al, (2010) [5]. This search was completed using the motif.find() function in the *bio3d* package in R [12]. Specifically, each amino acid sequence was searched for each of the 24 repeat motifs and the count of matches was reported back into the data. See Table 2. The breakdown of match frequencies by location is shown in Figure 4.

**Table 2:**
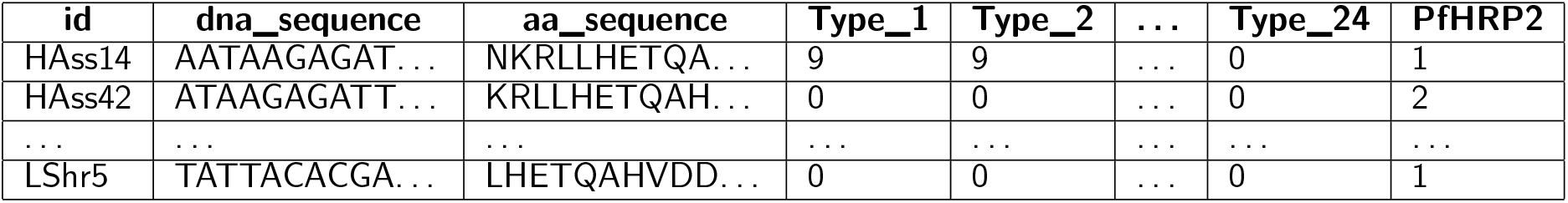
Example data format with counts of Types 1 through 24 matches in the amino acid sequence.

**Table 3:**
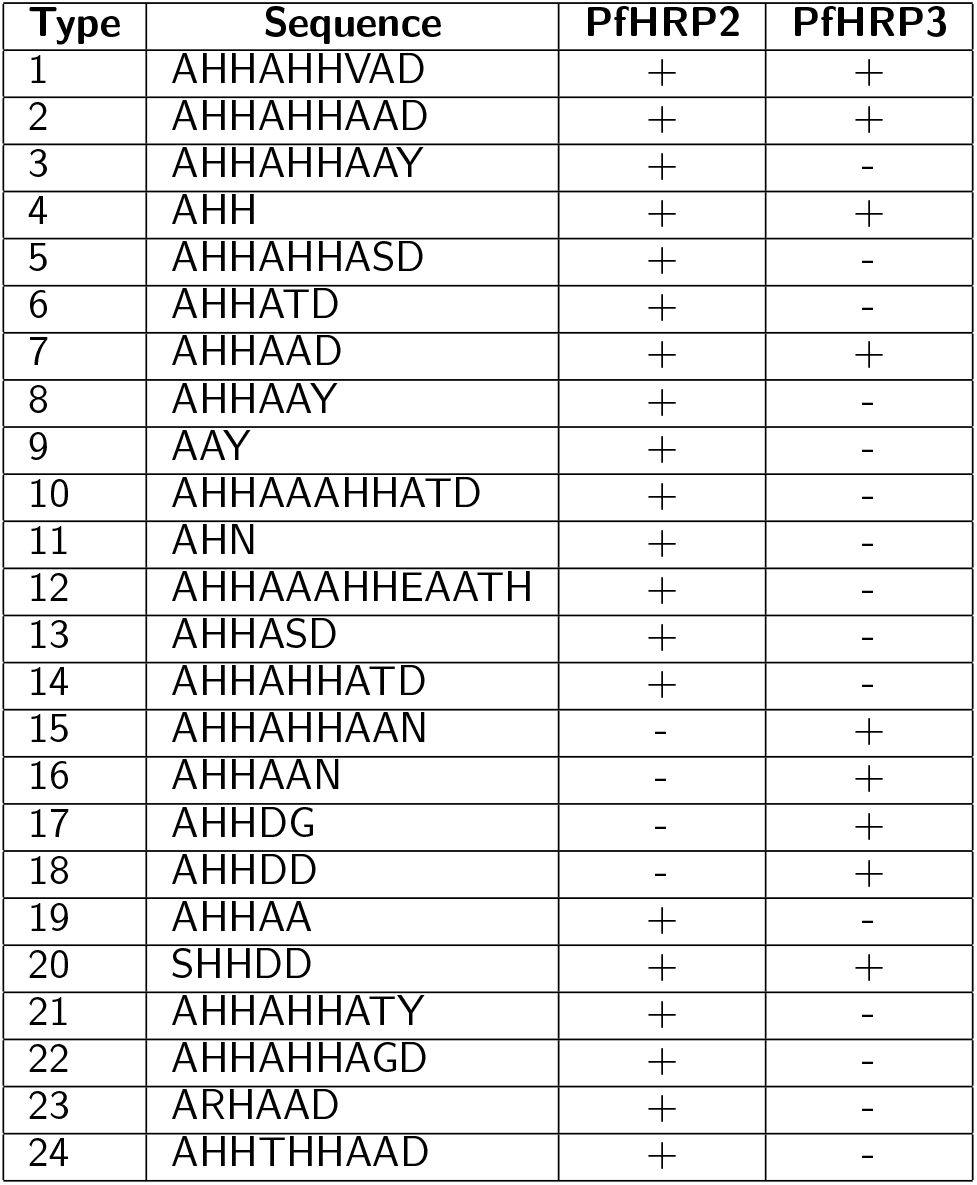
PfHRP2 and PFHRP3 repeat motif types as defined by Baker et al., 2010 [5].

**Figure 1:**
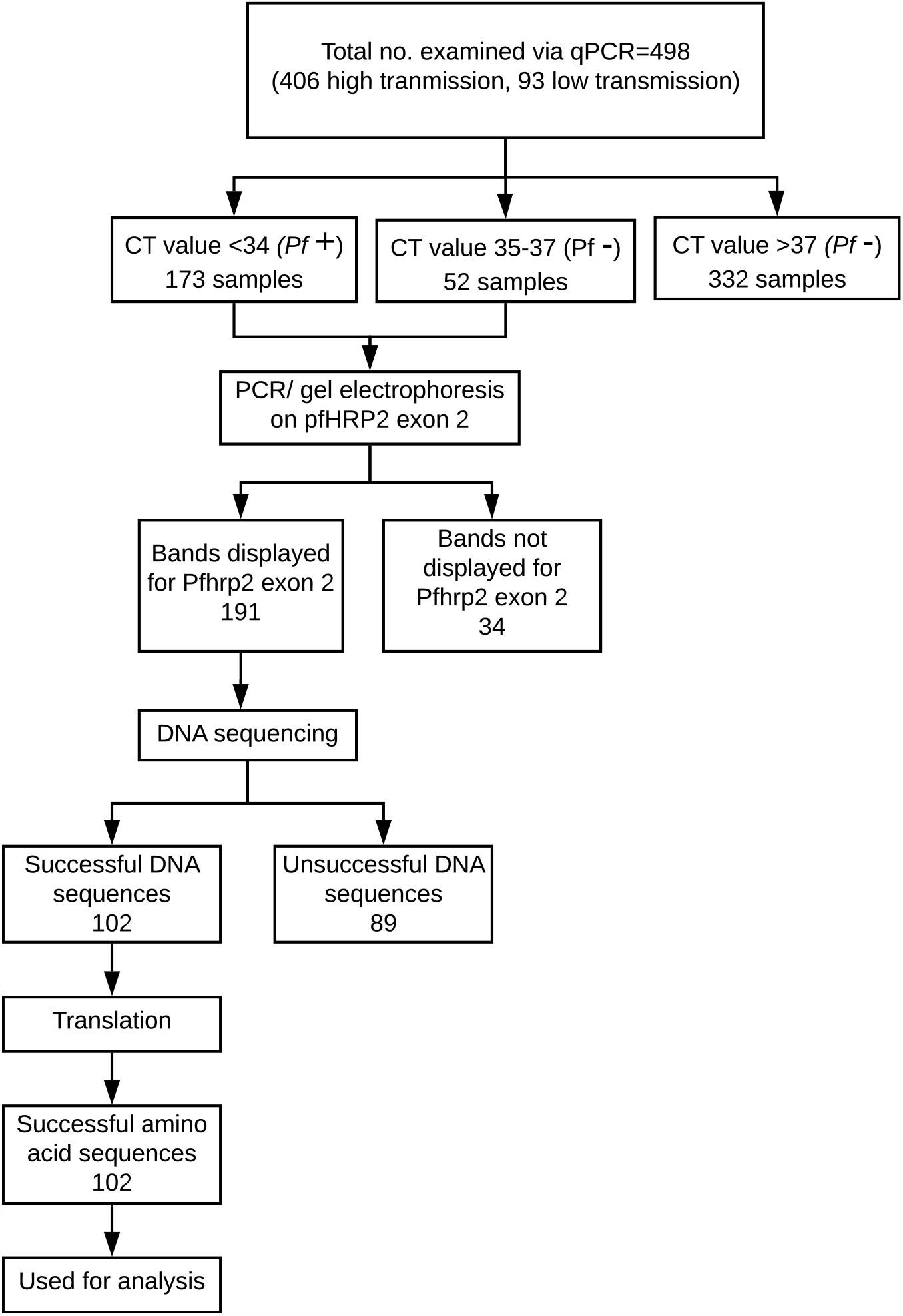
Breakdown of *P. falciparum* samples used in this study.

**Figure 2:**
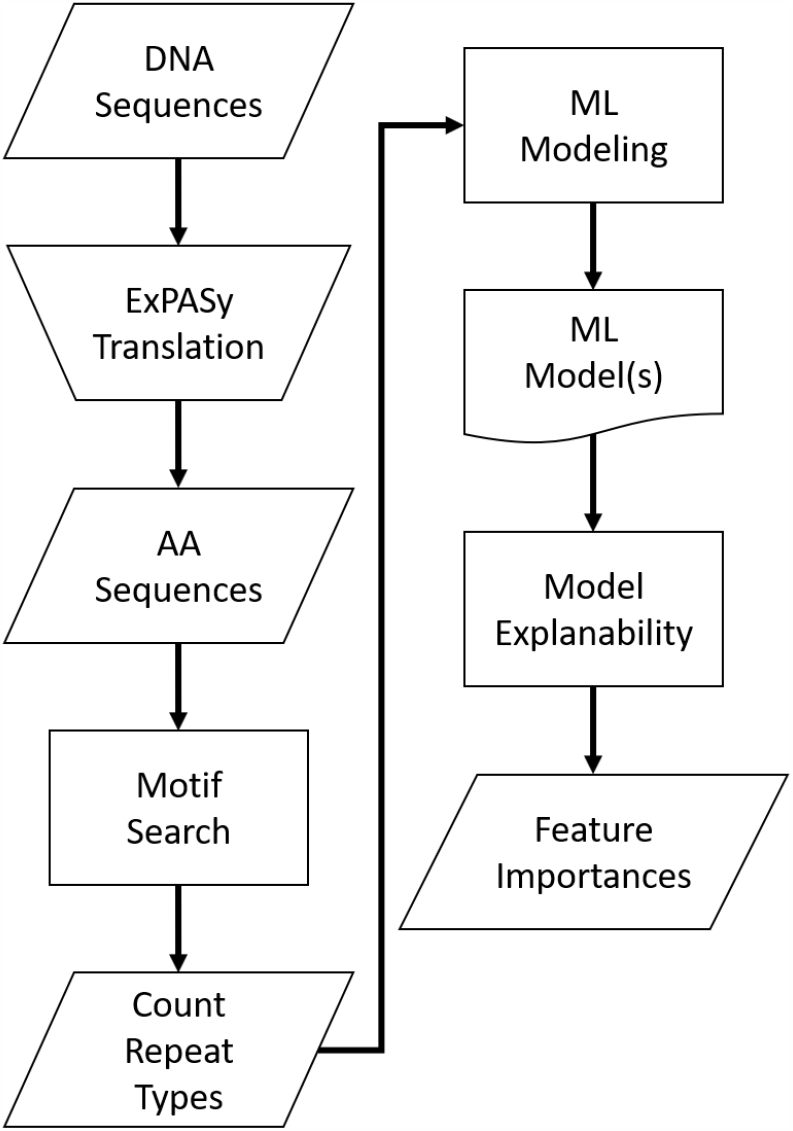
Analysis process flow.

**Figure 3:**
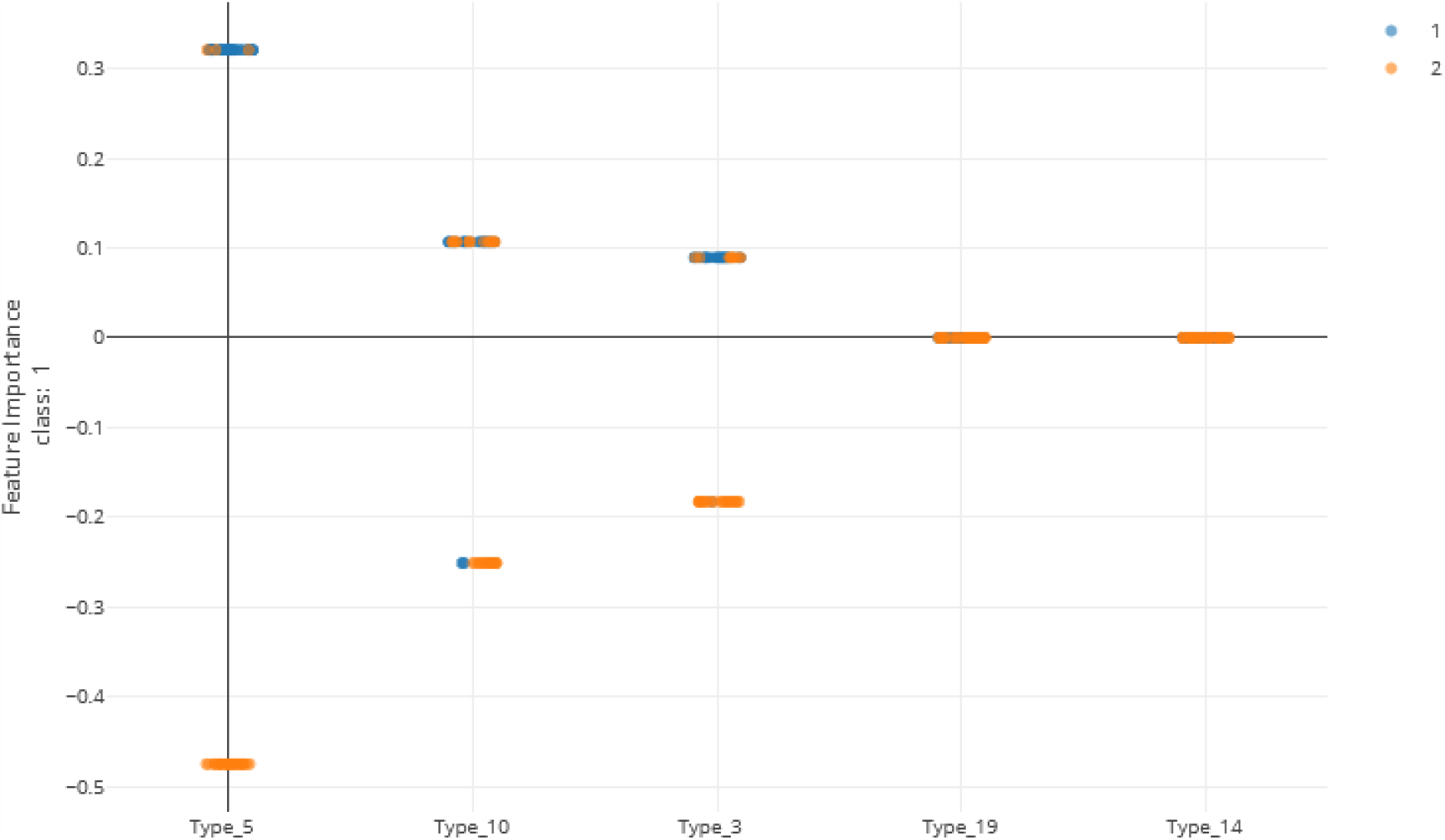
Local feature importance of the top 5 features. Note that only the top 3 have non-zero importances from the Voting Ensemble model using Types 1 through 24. Class “1” (orange dots) represents positive cases and class “2” (blue dots) represents negative cases of malaria.

**Figure 4:**
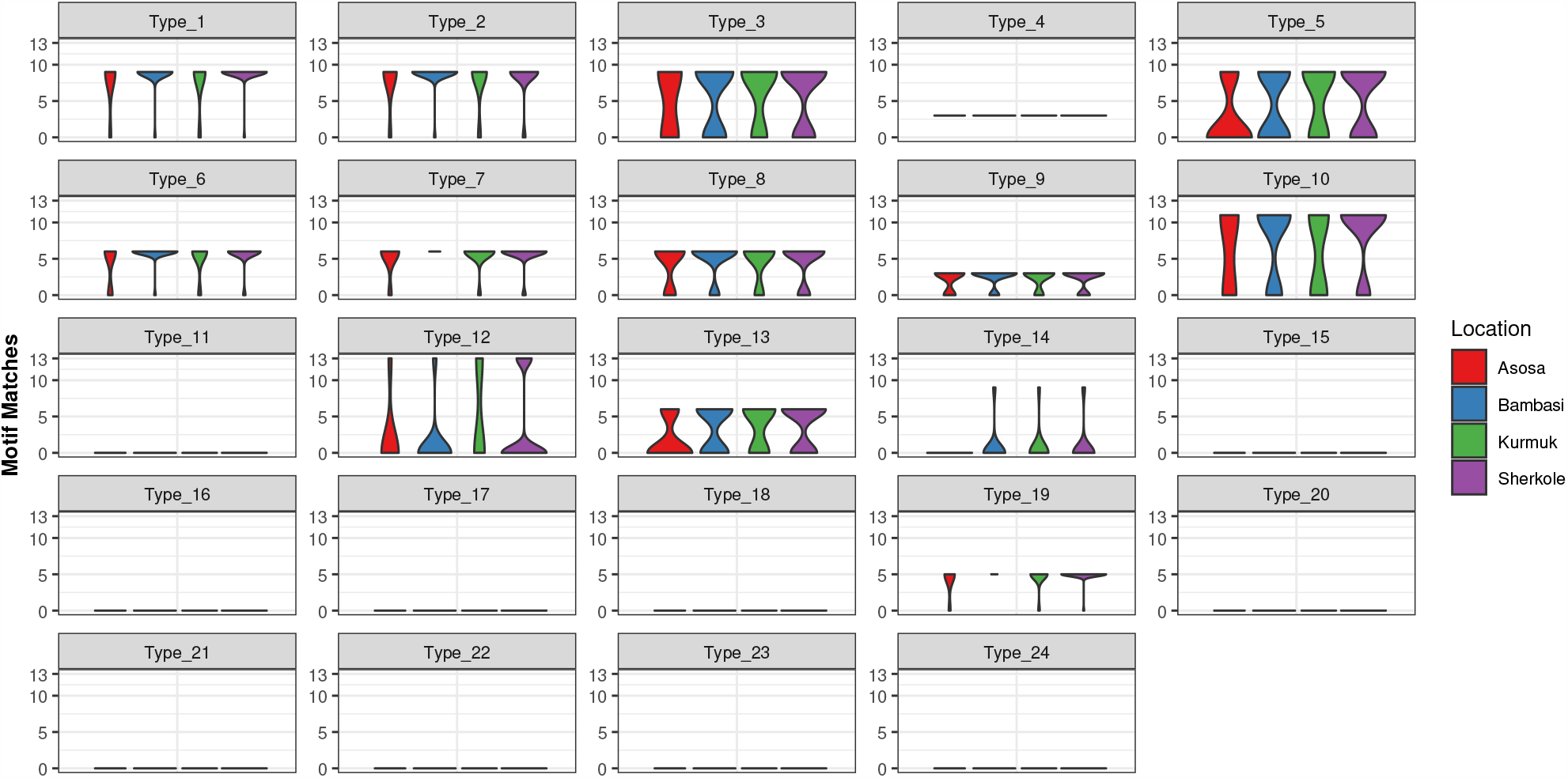
Type Frequencies by Location.

### 2.3 Machine Learning

In this work, three machine learning experiments were created on different sets of features: 1.) using only the types that are in the original Baker model (Types 2 and 7), 2.) using all motif repeat type counts (Types 1 through 24), and 3.) using only the features found to be important in the experiment with all motif repeat types (Types 3, 5, and 10). Note that the *PfHRP2* column in Table 2 is treated as the dependent variable in which a “1” represents a positive case of malaria and a “2” represents a negative case of malaria.

We used the Microsoft Azure Machine Learning Service [13] as the tracking platform for retaining model performance metrics as the various models were generated. For this use case, multiple machine learning models were trained using various scaling techniques and algorithms. Scaling and normalization methods are shown in Table 5. We then created two ensemble models of the individual models using stack ensemble and voting ensemble methods.

**Table 4:** Parameter settings for the model searches.

| Parameter | Value |
| --- | --- |
| Task | Classification |
| Training Time (hours) | 3 |
| Primary Metric | Precision score weighted |
| Validation type | Monte Carlo cross validation |

**Table 5:** Scaling function options in the machine learning model search [17].

| Scaling and Normalization | Description |
| --- | --- |
| StandardScaleWrapper | Standardize features by removing the mean and scaling to unit variance |
| MinMaxScaler | Transforms features by scaling each feature by that column's minimum and maximum |
| MaxAbsScaler | Scale each feature by its maximum absolute value |
| RobustScaler | This scales features by their quantile range |
| PCA | Linear dimensionality reduction using singular value decomposition of the data to project it to a lower dimensional space |
| TruncatedSVDWrapper | This transformer performs linear dimensionality reduction by means of truncated singular value decomposition. Contrary to PCA, this estimator does not center the data before computing the singular value decomposition. This means it can efficiently work with sparse matrices. |
| SparseNormalizer | Each sample (each record of the data) with at least one non-zero component is re-scaled independently of other samples so that its norm (L1 or L2) equals one |

The Microsoft AutoML package [14] allows for the parallel creation and testing of various models, fitting based on a primary metric. For this use case, models were trained using Decision Tree, Elastic Net, Extreme Random Tree, Gradient Boosting, Lasso Lars, LightGBM, RandomForest, and Stochastic Gradient Decent algorithms along with various scaling methods from Maximum Absolute Scaler, Min/Max Scaler, Principal Component Analysis, Robust Scaler, Sparse Normalizer, Standard Scale Wrapper, Truncated Singular Value Decomposition Wrapper (as defined in Table 5). All of the machine learning algorithms are from the *scikit-learn* package [15] except for LightGBM, which is from the *LightGBM* package [16]. The settings for the model sweep are defined in Table 4.

For the experiment using only Types 2 and 7, 35 models were trained. For the experiment using Types 1 through 24, 35 models were trained. For the experiments using Types 3, 5, and 10, 31 models were trained.

Once the individual models were trained, two ensemble models (voting ensemble and stack ensemble) were then created and tested for each experiment. The voting ensemble method makes a prediction based on the weighted average of the previous models’ predicted classification outputs whereas the stacking ensemble method combines the previous models and trains a meta-model using the elastic net algorithm based on the output from the previous models. The model selection method used was the Caruana ensemble selection algorithm [18].

## 3 Results

Metrics from the three experiments’ machine learning models (one each for the best ensemble model and a best singular model) are reported in Table 6. The precision-recall curves for these models are shown in Table 8 and the receiver operating characteristic (ROC) curves are shown in Table 7. The ideal scenario is shown as a dash-dot-dash (-.-) line. The best model overall is the Extreme Random Trees model using only Types 3, 5, and 10. This was determined by looking at the overall model metrics and the generated curves. Note that many models were generated for each experiment, some of which has equal overall performed. All model runs can be found in the Supplementary Data.

**Table 6:**
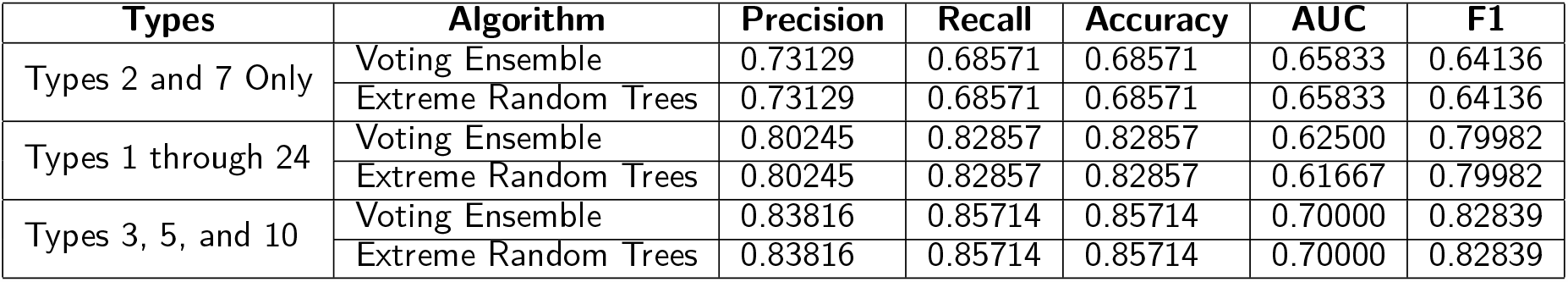
Model metrics for the best singular model and voting ensemble model for each experiment.

| Types | Algorithm | Precision | Recall | Accuracy | AUC | F1 |
| --- | --- | --- | --- | --- | --- | --- |
| Types 2 and 7 Only | Voting Ensemble | 0.73129 | 0.68571 | 0.68571 | 0.65833 | 0.64136 |
|  | Extreme Random Trees | 0.73129 | 0.68571 | 0.68571 | 0.65833 | 0.64136 |
| Types 1 through 24 | Voting Ensemble | 0.80245 | 0.82857 | 0.82857 | 0.62500 | 0.79982 |
|  | Extreme Random Trees | 0.80245 | 0.82857 | 0.82857 | 0.61667 | 0.79982 |
| Types 3, 5, and 10 | Voting Ensemble | 0.83816 | 0.85714 | 0.85714 | 0.70000 | 0.82839 |
|  | Extreme Random Trees | 0.83816 | 0.85714 | 0.85714 | 0.70000 | 0.82839 |

**Table 7:**
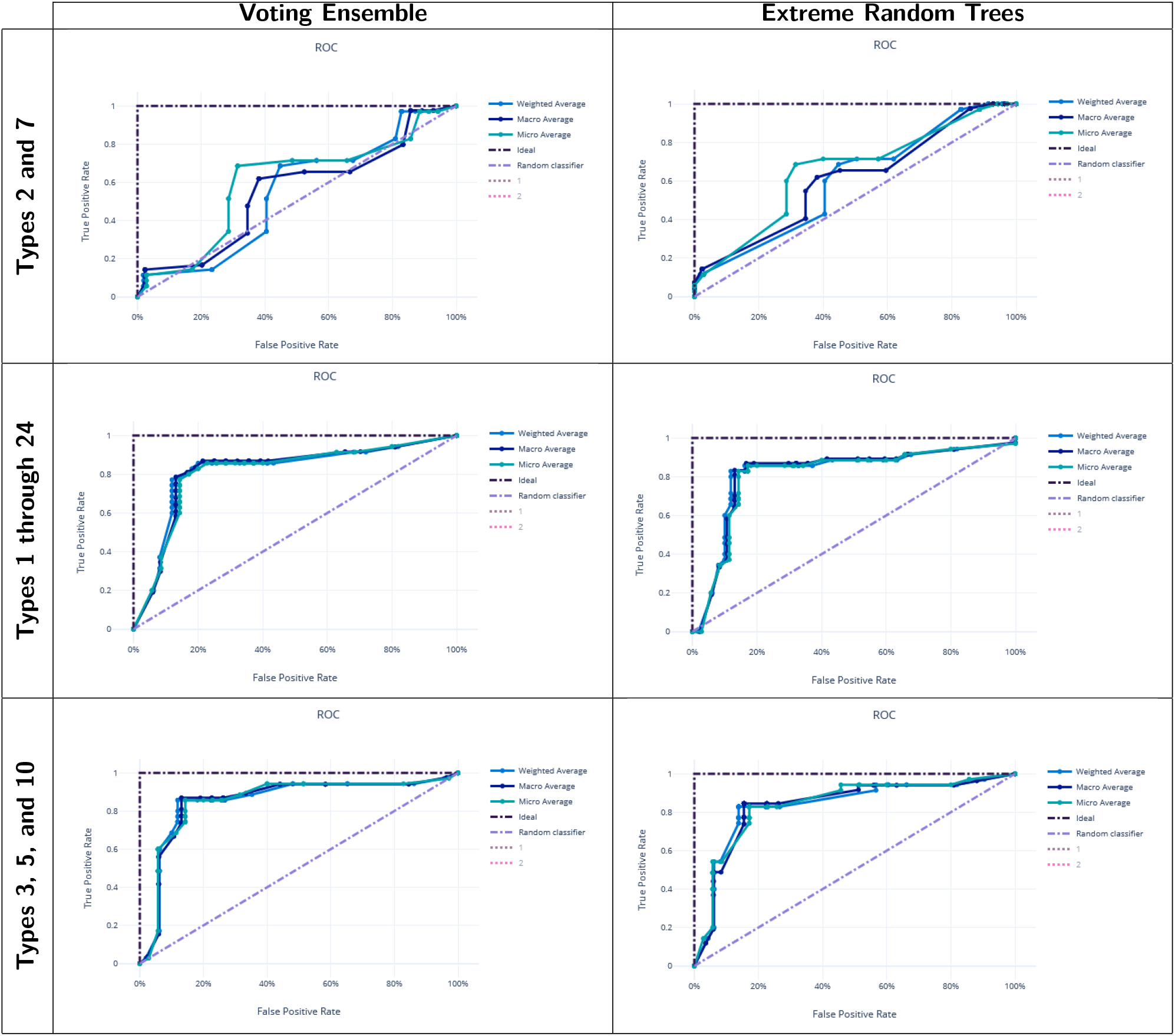
ROC Curves for the best singular model and voting ensemble model for each experiment.

**Table 8:**
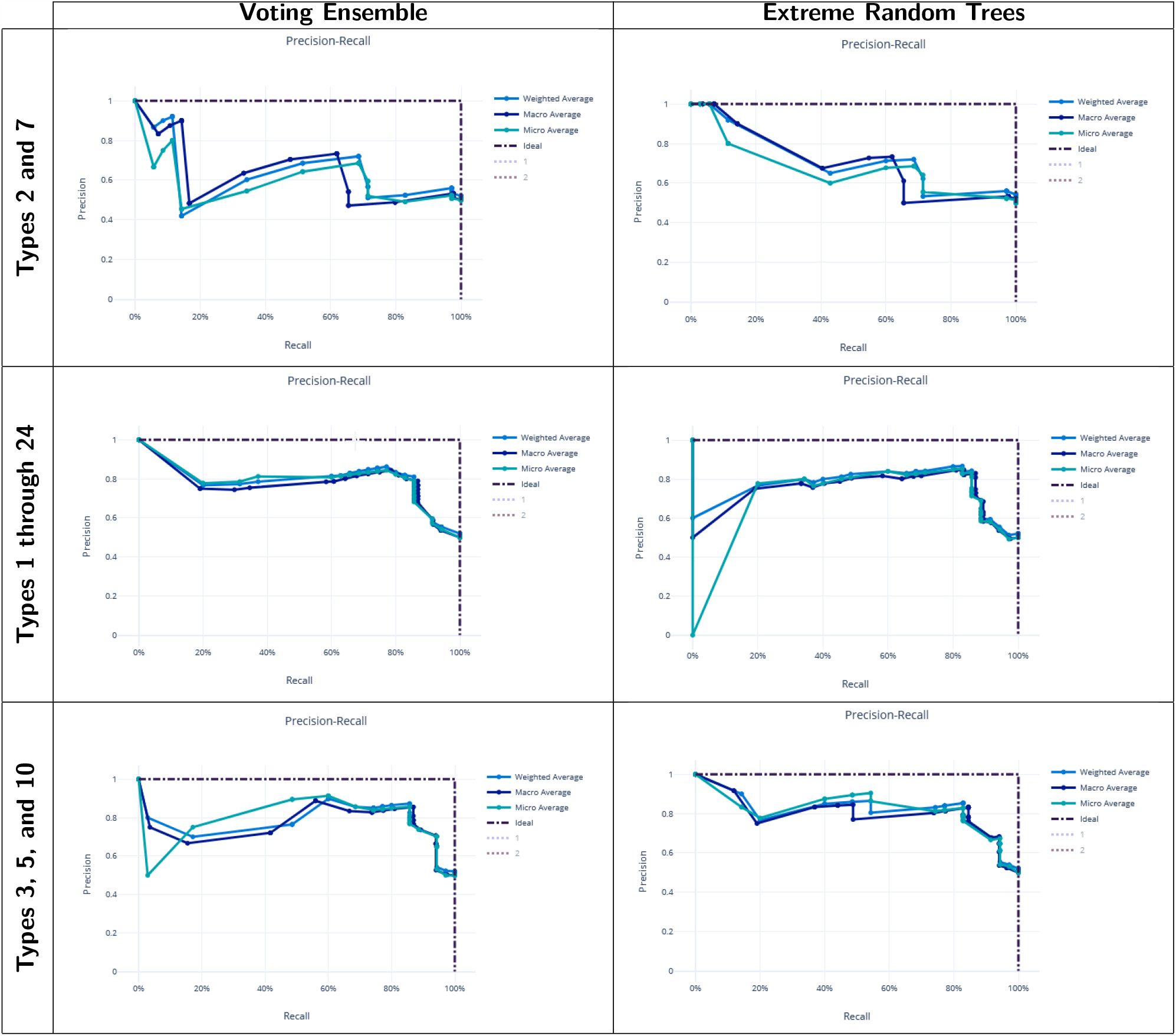
Precision-Recall Curves for the best singular model and voting ensemble model for each experiment.

### Feature importance

Feature importances were calculated using mimic-based model explanation of the voting ensemble model for Types 1 through 24. The mimic explainer works by training global surrogate models to mimic blackbox model [19]. The surrogate model is an interpretable model, trained to approximate the predictions of a black box model as accurately as possible [20].

In the Voting Ensemble model using Types 1 through 24, Types 3, 5, and 10 were found to have non-zero importance. See Figure 3.

### 3.1 Repeat Type Prevalence

As shown in Figure 4 and Table 10, many of the repeat types described by Baker et al., 2010 [5] (Table 3) are represented in the Ethiopian sequences analyzed in this study. Specifically, Types 1-10, 12-14, and 19 were found among these isolates. This is in general agreement to a similar report by Willie et al., 2018 [21] using samples collected from Papua New Guinea. They report that Types 1, 2, 6, 7, and 12 were present in almost all (*≥* 89%) sequences, Types 3, 5, 8, and 10 were present in most (*≥* 56%) sequences, and Type 4, 13, and 19 were seen in ≤ 33% of sequences. In contrast, we see a higher prevalence of Types 4 and 19 and a lower prevalence of Type 12 than in the previous study.

**Table 9:** Global and local feature importances of all features with non-zero importance (Types 3, 5, and 10) from the Voting Ensemble model using Types 1 through 24.

|  | Global Importance | Local Importance |
| --- | --- | --- |
| **Type 3** | 0.15547 | Min: -0.22644<br>Average: -4.14E-19<br>Std. Dev: 0.16433<br>Max: 0.22644 |
| **Type 5** | 0.48787 | Min: -0.60532<br>Average: -1.66E-18<br>Std. Dev: 0.49919<br>Max: 0.60533 |
| **Type 10** | 0.28736 | Min: -0.48132<br>Average: -2.49E-18<br>Std. Dev: 0.31516<br>Max: 0.48132 |

**Table 10:** Overall prevalence of each repeat type by location. Values represent the percentage of samples in which the repeat type was found.

| Type | Asosa | Bambasi | Kurmuk | Sherkole | Overall |
| --- | --- | --- | --- | --- | --- |
| 1 | 85.71% | 97.06% | 84.62% | 97.92% | 95.10% |
| 2 | 85.71% | 97.06% | 84.62% | 91.67% | 92.16% |
| 3 | 57.14% | 61.76% | 69.23% | 66.67% | 64.71% |
| 4 | 100.00% | 100.00% | 100.00% | 100.00% | 100.00% |
| 5 | 28.57% | 50.00% | 61.54% | 62.50% | 55.88% |
| 6 | 71.43% | 97.06% | 84.62% | 93.75% | 92.16% |
| 7 | 85.71% | 100.00% | 92.31% | 93.75% | 95.10% |
| 8 | 71.43% | 82.35% | 76.92% | 77.08% | 78.43% |
| 9 | 71.43% | 82.35% | 76.92% | 77.08% | 78.43% |
| 10 | 57.14% | 67.65% | 53.85% | 77.08% | 69.61% |
| 11 | 0.00% | 0.00% | 0.00% | 0.00% | 0.00% |
| 12 | 14.29% | 8.82% | 38.46% | 25.00% | 20.59% |
| 13 | 28.57% | 55.88% | 61.54% | 62.50% | 57.84% |
| 14 | 0.00% | 8.82% | 7.69% | 10.42% | 8.82% |
| 15 | 0.00% | 0.00% | 0.00% | 0.00% | 0.00% |
| 16 | 0.00% | 0.00% | 0.00% | 0.00% | 0.00% |
| 17 | 0.00% | 0.00% | 0.00% | 0.00% | 0.00% |
| 18 | 0.00% | 0.00% | 0.00% | 0.00% | 0.00% |
| 19 | 85.71% | 100.00% | 92.31% | 97.92% | 97.06% |
| 20 | 0.00% | 0.00% | 0.00% | 0.00% | 0.00% |
| 21 | 0.00% | 0.00% | 0.00% | 0.00% | 0.00% |
| 22 | 0.00% | 0.00% | 0.00% | 0.00% | 0.00% |
| 23 | 0.00% | 0.00% | 0.00% | 0.00% | 0.00% |
| 24 | 0.00% | 0.00% | 0.00% | 0.00% | 0.00% |

In another study by Bharti et al., 2016 [22] that used samples collected from multiple sites in India reported that Types 1, 2, 7, and 12 were seen in 100% of their sequences. However, in our sequences from Ethiopia, we see multiple examples where these repeats are not present, especially Type 12.

## 4 Discussion

Here we show the utility of machine learning in the identification of important factors in malaria diagnosis. Previous modeling by Baker et al., 2005 [1] had shown that the parasitic infection can be diagnosed by looking at the prevalence of particular types of amino acid repeats. The original regression-based model is no longer valid and, in this study, we show that the modeling of Types 2 and 7 using more sophisticated machine learning algorithms fail to produce a reliable model. However, the usage of Types 1 through 24 proves to make effective models that better characterize test performance to detect *P. falciparum* infections in our dataset. Furthermore, the usage of machine learning model explanability helps to pinpoint particular features of interest. In this case, Types 3, 5 and 10 reveal better diagnostic sensitivity for these malaria isolates collected from regions of Ethiopia.

Several studies have indicated that the Type 2 repeat (AHHAHHAAD) and Type 7 repeat (AHHAAD) have been described as possible epitopes targeted by monoclonal antibodies used to detect PfHRP2 [5, 23]. The highest frequency Types 2, 4, and 7 are also observed in some African countries [24]. This is in agreement with our findings in this work for the Types that have a high prevalence frequency (between 85%-100%). However, our analysis here may reveal better diagnostic sensitivity for Types 3, 5, and 10, which have lower frequencies (between ∼ 28%-70%) among the malaria isolates collected from our study area in Ethiopia. These Type prevalences by region are shown in Table 10.

This work posits the idea that RDTs can be revised to accommodate the genetic differences seen in today’s *P. falciparum* infections and malaria cases. Future versions of RDTs may be improved using our novel methodology for identifying genetic variants to improve test sensitivity. Though more work is to be done to empirically validate these findings, this *in silico* simulation may direct where to take experimental testing next. Also, while this work showcases important histidine-rich repeats of Types 3, 5, and 10, this is specific to the Ethiopian sequences used in this study and other *P. falciparum* strains in other regions may result in different results. Furthermore, training machine learning models on sets of malaria sequences from other areas such as Papua New Guinea, India, or other areas of Africa may reveal that different repeats are important in those areas, likely suggesting the RDTs may need to be region-specific due to variations in *P. falciparum* across the globe.

## Data Availability

All data, scripts, and model outputs are hosted on GitHub at: https://github.com/colbyford/pfHRP_MLModel

## 5 Supplementary Materials

All data, scripts, and model outputs are hosted on GitHub at: github.com/colbyford/pfHRP_MLModel

## 6 Author Contributions

G.A. and L.G. designed and performed the patient recruitment and sampling. G.A, L.G. and D.J. managed ethical approval, funding, and visas. G.A., K.L., K.B., and C.C.D performed the DNA extractions, RT-PCR, PCR, and sequencing of the samples under the direction of L.G., D.J., and E.L. K.B. and D.J. performed the DNA to amino acid translations. C.T.F. performed the motif search for repeat types and performed all the machine learning and model interpretability work. All authors reviewed this manuscript.

## 7 Acknowledgements

We thank the family of Carol Grotnes Belk for financial support. We acknowledge the administrative, salary, and laboratory support of these entities at the University if North Carolina at Charlotte: the Office of International Student Scholars, the Bioinformatics Research Center, the College of Computing and Informatics, the Department of Bioinformatics and Genomics, the College of Liberal Arts and Sciences, and the Department of Biological Sciences. The field data collection portion of this work was funded in part by Addis Ababa University Thematic Research.

## 9 Ethics Statement

Scientific and ethical clearance was obtained from the Institutional Scientific and Ethical Review Boards of Addis Ababa University in Ethiopia and The University of North Carolina, Charlotte, USA. Written informed consent/assent for study participation was obtained from all consenting heads of households, parents/guardians (for minors under age of 18), and each individual who was willing to participate in the study.

## References

[1] Baker, J. et al.. Genetic Diversity of Plasmodium falciparum Histidine-Rich Protein 2 (PfHRP2) and Its Effect on the Performance of PfHRP2-Based Rapid Diagnostic Tests. The Journal of Infectious Diseases 192, 3870–877 (2005). URL https://doi.org/10.1086/432010. https://academic.oup.com/jid/article-pdf/192/5/870/2404536/192-5-870.pdf.

[2] Organization, W. H. Fact sheet about malaria. URL https://www.who.int/news-room/fact-sheets/detail/malaria.

[3] How malaria rdts work (2015). URL https://www.who.int/malaria/areas/diagnosis/rapid-diagnostic-tests/about-rdt/en/.

[4] Ouattara, A. et al.. Polymorphisms in the k13-propeller gene in artemisinin-susceptible plasmodium falciparum parasites from bougoula-hameau and bandiagara, mali. The American Journal of Tropical Medicine and Hygiene 92, 1202–1206 (2015). URL http://www.ajtmh.org/content/journals/10.4269/ajtmh.14-0605http://www.ajtmh.org/content/journals/10.4269/ajtmh.14-0605

[5] Baker, J. et al. Global sequence variation in the histidine-rich proteins 2 and 3 of plasmodium falciparum: implications for the performance of malaria rapid diagnostic tests. Malaria Journal 9, 129 (2010). URL https://doi.org/10.1186/1475-2875-9-129.

[6] Davis, S. et al. Leveraging crowdsourcing to accelerate global health solutions. Nature Biotechnology 37, 848–850 (2019). URL https://doi.org/10.1038/s41587-019-0180-5.

[7] Ford, C. T. & Janies, D. Ensemble machine learning modeling for the prediction of artemisinin resistance in malaria. F1000Research 9 (2020).

[8] R.B., M., R.J., C., F., S. & M.C., S.-M. Evaluation of three different dna extraction methods from blood samples collected in dried filter paper in plasmodium subpatent infections from the amazon region in brazil. Revista do Instituto de Medicina Tropical de Sao Paulo 55, 205–208 (2013).

[9] G., S. et al. High sensitivity of detection of human malaria parasites by the use of nested polymerase chain reaction. Molecular and Biochemical Parasitology 61, 315–320 (1993).

[10] Alemayehu, G. S. et al. Evaluation of PfHRP2 and PfLDH Malaria Rapid Diagnostic Test Performance in Assosa Zone, Ethiopia. BMC Infectious Diseases, In Review. (2020).

[11] Parr, J. B., Anderson, O., Juliano, J. J. & Meshnick, S. R. Streamlined, pcr-based testing for pfhrp2- and pfhrp3-negative plasmodium falciparum. Malaria Journal 17, 137 (2018). URL https://doi.org/10.1186/s12936-018-2287-4.

[12] B.J., G., A.P.C., R., K.M., E., J.A., M. & L.S.D., C. Bio3d: An r package for the comparative analysis of protein structures. Bioinformatics 22, 2695–2696 (2006).

[13] Microsoft Azure Machine Learning Service (2019). URL https://azure.microsoft.com/en-us/services/machine-learning/.

[14] Microsoft. Azure Machine Learning AutoML Core version 1.0.79 (2019). URL https://pypi.org/project/azureml-automl-core/.

[15] Pedregosa, F. et al. Scikit-learn: Machine learning in Python. Journal of Machine Learning Research 12, 2825–2830 (2011).

[16] Ke, G. et al. Lightgbm: A highly efficient gradient boosting decision tree. In Guyon, I. et al. (eds.) Advances in Neural Information Processing Systems 30, 3146–3154 (Curran Associates, Inc., 2017). URL http://papers.nips.cc/paper/6907-lightgbm-a-highly-efficient-gradient-boosting-decision-tree.pdf.

[17] Microsoft. Microsoft Azure Machine Learning - AutoML Preprocessing (2019). URL https://docs.microsoft.com/en-us/azure/machine-learning/concept-automated-ml#automatic-preprocessing-standard.

[18] Caruana, R., Niculescu-Mizil, A., Crew, G. & Ksikes, A. Ensemble selection from libraries of models. In Proceedings of the Twenty-first International Conference on Machine Learning, ICML’04, 18– (ACM, New York, NY, USA, 2004). URL http://doi.acm.org/10.1145/1015330.1015432.

[19] Lundberg, S. M. & Lee, S.-I. A unified approach to interpreting model predictions. In Guyon, I. et al. (eds.) Advances in Neural Information Processing Systems 30, 4765–4774 (Curran Associates, Inc., 2017). URL http://papers.nips.cc/paper/7062-a-unified-approach-to-interpreting-model-predictions.pdf.

[20] Molnar, C. Interpretable Machine Learning (2019). https://christophm.github.io/interpretable-ml-book/.

[21] Willie, N., Zimmerman, P. A. & Mehlotra, R. K. Plasmodium falciparum histidine-rich protein 2 gene variation in a malaria-endemic area of papua new guinea. The American Journal of Tropical Medicine and Hygiene 99, 697–703 (2018). URL https://www.ajtmh.org/content/journals/10.4269/ajtmh.18-0137.

[22] Bharti, P. K. et al. Prevalence of pfhrp2 and/or pfhrp3 gene deletion in plasmodium falciparum population in eight highly endemic states in india. PLOS ONE 11, 1–16 (2016). URL https://doi.org/10.1371/journal.pone.0157949.

[23] Lee, N. et al. Identification of optimal epitopes for Plasmodium falciparum rapid diagnostic tests that target histidine-rich proteins 2 and 3. J. Clin. Microbiol. 50, 1397–1405 (2012).

[24] Deme, A. B. et al. Analysis of pfhrp2 genetic diversity in senegal and implications for use of rapid diagnostic tests. Malaria Journal 13, 34 (2014). URL https://doi.org/10.1186/1475-2875-13-34.

